# Cancer epidemiology and survival in a high HIV prevalence region of Kenya: a 10-year retrospective cohort study

**DOI:** 10.64898/2026.09.28.26364183

**Authors:** Dorothy Mangale, Harriet Fridah Adhiambo, Phiona Adagi, Donella Atieno, Benjamin Bowe, Caroline Wafula, Angela McLigeyo, Thomas A. Odeny

## Abstract

**Background:** Cancer incidence and mortality are rising in Africa, yet the distribution and determinants of cancer in high HIV-prevalence regions remain limited. We characterized cancer epidemiology, treatment, and survival among adults in western Kenya, where HIV prevalence ranges between 10.3% and 11.7%.

**Methods:** This retrospective cohort study included adults (≥18 years) with cancer treated at Jaramogi Oginga Odinga Teaching and Referral Hospital (JOOTRH) in Kisumu, Kenya, between Jan 1, 2014, and Dec 31, 2024, followed through June 30, 2025. Data were abstracted from paper records into REDCap. Negative binomial models estimated annual trends, Kaplan-Meier curves evaluated survival, and Cox proportional hazards models evaluated mortality

**Findings:** Among 3978 patients (64% female; median age 53.9 years, IQR (42.0, 65.1), annual case counts rose from 29 to 708, averaging a 38% annual increase (95% CI 29-48). Cervical (24%), esophageal (13.6%), breast (12%), prostate (8%), and colorectal (4%) cancers accounted for 62% of cases. 1305 (33%) of cancer patients were living with HIV and 802 (20%) had unknown HIV status. Among patients whose cancer was assigned a stage, 72% presented at stage III-IV. 1973 (50%) received cancer-directed treatment, and 51% had an unscheduled clinic visit lapse of at least 180 days. Cancer-directed treatment was the strongest protective factor for mortality (adjusted hazard ratio [aHR] 0·27, 95% CI 0·22-0·32); HIV-positive status (aHR 1·43, 1·19-1·71), HIV-unknown status (aHR 1·48, 1·20-1·81), and poor or missing performance status were independently associated with higher mortality, robust across sensitivity analyses.

**Interpretation:** Cancer mortality was strongly associated with treatment and HIV status at diagnosis. Closing the treatment-access gap and adopting routine opt-out HIV testing at the first oncology visit are essential for integrated cancer care in high-HIV-prevalence settings.

**Funding:** Research reported in this publication was supported by the American Cancer Society; the Washington University in St. Louis Institute of Clinical and Translational Sciences through a grant from the National Center for Advancing Translational Sciences (NCATs) of the National Institutes of Health; Just-in-Time funding, and the National Cancer Institute Center Support Grant P30CA091842.

## INTRODUCTION

Cancer incidence and mortality are rising disproportionately in low- and middle-income countries (LMICs).^1^ Between 2024 and 2050, cancer deaths are projected to rise by about 91% in LMIC, more than double the 43% increase forecast for high-income countries.^2^ In Africa, annual cancer mortality is projected to nearly double from approximately 520,348 in 2020 to about 1 million by 2030.^3^ In Kenya, GLOBOCAN 2022 estimated 44,726 new cancer cases and 29,317 deaths, with breast, cervical, prostate, esophageal, and colorectal cancer ranking among the leading cancers nationally.^4^ However, nationally aggregated estimates may not adequately capture geographic and regional heterogeneity in cancer burden across the country. ^5^ Yet, these estimates are derived from limited registry data and can obscure important regional variation in epidemiology, especially when infectious, environmental, and behavioral risk factors differ from national averages.

This gap is particularly pronounced in western Kenya, where HIV prevalence remains high, reaching 11.7% in Homabay County compared with 3.2% nationally among persons aged 15-49 years^6^ and with distinct exposure to oncogenic viruses such as Epstein-Barr virus (EBV), Kaposi sarcoma-associated herpesvirus (KSHV), and human papillomavirus (HPV).^7–10^ Consequently, this region faces a dual burden of virus-associated malignancies alongside a growing incidence of non-communicable cancers. Although breast and cervical cancers in women and prostate cancer in men predominate across Africa,^11^ older data from 1999-2006 in Kenya, at the peak of the HIV epidemic, identified esophageal squamous cell cancer (ESCC) as a leading malignancy, alongside prostate cancer, and a persistent burden of HIV-associated (previously referred to as AIDS-defining) malignancies such as Kaposi sarcoma, cervical cancer, and non-Hodgkin lymphoma, despite scale up of antiretroviral therapy.^12^ Additional patterns, including high cervical cancer rates, rising colorectal cancer incidence, and regionally elevated stomach cancer rates, suggest important geographic and etiologic heterogeneity.^13^ However, the extent to which region-specific risk factors translate into observable differences in cancer distribution and outcomes remains understudied. Importantly, although cancer patterns among people with HIV (PWH) have been characterized in multiple settings, including South Africa,^14^ Uganda,^15^ Malawi,^16^ and Botswana, ^17^ using registry-based and clinical cohort studies, the clinical and prognostic significance of unknown HIV status among cancer patients remains largely unaddressed, and comparable data from the Lake Victoria basin, a recognized HIV epicenter with distinct environmental and behavioral exposures, are lacking.

To address this gap, we conducted a retrospective cohort study of adults receiving cancer care at Jaramogi Oginga Odinga Teaching and Referral Hospital (JOOTRH), a major tertiary referral center in western Kenya. Using 10 years of routinely collected clinical data, we describe the distribution of cancer diagnoses, treatment patterns, and survival outcomes, and examine the contribution of HIV status to survival. To our knowledge, this represents the first comprehensive assessment of cancer in this high-HIV-prevalence region.

Panel 1: Research in context

**Evidence before this study**

Cancer incidence and mortality are rising faster in Africa than in any other region, but national registry estimates obscure substantial geographic heterogeneity in cancer type, risk factors, and outcomes. Cohort and registry-linkage studies from Uganda, Malawi, Botswana, and South Africa have shown that people with HIV carry a disproportionate burden of Kaposi sarcoma, cervical cancer, and non-Hodgkin lymphoma, and generally have worse cancer survival than HIV-negative patients. Separate work on HIV care delivery in East Africa, including patient navigation, phone-based tracing, and community health worker linkage, has shown that gaps in care engagement are addressable with active, low-cost strategies. To our knowledge, no prior study has characterized decade-long trends in cancer incidence, treatment uptake, and survival at a referral hospital in the Lake Victoria basin, a region with a disproportionately high HIV burden compared with the national prevalence of 3.2%. Nor has any prior cancer cohort in the region examined survival among patients with unknown HIV status as its own category, despite this being a common feature among newly diagnosed cancer patients.

**Added value of this study**

Using 10 years of clinical data abstracted from Jaramogi Oginga Odinga Teaching and Referral Hospital (JOOTRH), the referral center for over 10 million people in western Kenya, we show that oncology caseload rose roughly 23-fold over the study period, with cervical and prostate cancer increasing fastest. Nearly a third of patients (33%) were living with HIV, well above regional background prevalence, and HIV positivity reached 82% among patients with Kaposi sarcoma and 85% among patients with penile cancer. Patients with unknown HIV status had the worst survival of any group (adjusted HR 1.48), while those with confirmed HIV reported an adjusted HR of 1.43. This suggests that missing HIV status marks poor engagement with care and likely undiagnosed HIV rather than a gap in documentation.

**Implications of all the available evidence**

Read alongside prior evidence on HIV-associated cancer risk and the demonstrated effectiveness of retention-focused HIV care interventions in East Africa, our findings support routine opt-out HIV testing at oncology intake, as well as adaptation of existing HIV retention strategies (such as patient navigation, community-based tracing) to reduce loss to follow-up in cancer care. As cancer caseloads at high-HIV-prevalence referral centers continue to grow, facility-level evidence will complement modeled projections and improve precision of estimates.

## METHODS

### Study design and setting

This is a retrospective cohort study of adults presenting for cancer care at JOOTRH oncology clinic in Kisumu County, western Kenya. JOOTRH is the cancer referral hospital for 14 counties around the Lake Victoria region, representing a population of over 10 million. The region bears a disproportionately high burden of HIV. The Kenya National Syndemic Disease Control Council 2026 report estimates the HIV prevalence among adults aged 15-49 years to range between 10.3% in Siaya County and 11.7% in both Kisumu and Homa Bay, against a national prevalence of 3.2%.^6^ Available cancer services include screening and prevention programs, chemotherapy, surgical oncology, and palliative care. During the study period, radiotherapy was unavailable at the site, and patients requiring this service were referred to facilities outside the region.

### Study population

We included adults aged 18 years or older presenting for cancer care at JOOTRH between January 1, 2014, and December 31, 2024, with follow-up through June 30, 2025. Eligible participants were required to have a confirmed histopathological cancer diagnosis, or, where histopathology was missing, to have documentation of receiving cancer-directed treatment.

### Data Sources and Data Collection

Data were retrospectively abstracted from paper-based oncology clinic registers, clinic logs, and individual patient clinical records, and entered in a standardized electronic data collection tool hosted on REDCap.^18^ The abstraction tool captured demographic, clinical, diagnostic, treatment, and outcome information.

Records were reviewed for duplicate entries, internal inconsistencies, and incomplete diagnostic information. For patients with discordant diagnoses across sources, missing histopathology, or incomplete records but clear documentation of cancer-directed care, final cancer classification was determined by review of available clinical documentation by an oncologist (FA, TAO).

### Variables

Key demographic variables included age, sex, marital status, education level, occupation, insurance status, and geographic identifiers (county, subcounty, location, sublocation, and village). Clinical variables included date of cancer diagnosis, cancer type (according to WHO classification), clinical stage, Eastern Cooperative Oncology Group (ECOG) performance status,^19^ tumor location, cancer treatment, and HIV status. Pathological characteristics included histology, tumor grade, and pathological staging. Additional administrative and health system variables, such as medical record numbers, laboratory identifiers, and accession numbers, were available to support data linkage and longitudinal tracking. Where available, HIV-related clinical data, including antiretroviral therapy regimen, CD4 T-cell counts, and viral load, were included.

### Outcomes

The main outcomes were overall case counts, overall HIV status, overall survival, survival by treatment, and factors associated with survival. Overall survival was defined as the time from cancer diagnosis to death from any cause. Death was the event of interest; patients who were lost to follow-up, transferred out, or remained alive at the end of follow-up (June 30, 2025) were censored at their last known date of follow-up. Loss to follow-up was defined as a lapse in clinic visits for ≥ 180 days.

### Statistical Analysis

Participant characteristics are described as mean (standard deviation), median (interquartile range), or frequency and proportions as appropriate. Differences in characteristics were examined using Mann-Whitney U tests for continuous variables, and Chi-squared or Fisher’s exact tests for categorical variables.

Changes in cancer cases over time were examined using a negative binomial model with a log link, and the average annual change in the number of cancers and the percentage change are provided with their 95% confidence intervals.

Mortality in the 5 years after diagnosis is visualized with Kaplan-Meier curves, and differences between curves are assessed with the Wilcoxon test. Risk factors for mortality were analyzed using Cox proportional hazards models, starting at the time of diagnosis and censored at the time of last follow-up, death, transfer out, or administrative end of follow-up (June 30, 2025). Participants who were recorded as dead but were missing a date of death had their date of death imputed as the midpoint between the last clinic date and the administrative end of follow-up. The proportional hazards assumptions were examined visually via log-negative log plots and evaluated through the inclusion of time interaction term in the models and were met. Unadjusted and adjusted hazard ratios (HR) with 95% confidence intervals (CI) are presented, and adjusted HR are visualized in a forest plot.

To examine the robustness of results, we conducted two sets of sensitivity analyses. First, to examine the impact of potential biases due to informative censoring from competing risks, we alternatively, at the time of censoring due to last follow-up or transfer out, assigned an outcome to occur or not to occur through the administrative end of follow-up.^20^ Second, we conducted sensitivity analysis assuming that 1) the last known alive date is the last clinic visit, making the last clinic visit the earliest possible date of death, and 2) death occurred at the end of the observation time (the latest possible date of death) (see Appendix 1, Figures A1-A3 for a visual summary).

All analyses were two-tailed, where a p-value <0.05 or a 95% CI not containing unity was considered statistically significant. Missing data was assigned to an unknown category for categorical variables. Analyses were done using SAS Enterprise Guide 8.3 (SAS Institute Inc., NC), and visualization was done in R 4.3.1 (R Foundation for Statistical Computing, Austria).^21^

## Data Availability

De-identified individual participant data from this study are available upon reasonable request to the corresponding author, subject to approval by the Kenya Medical Research Institute (KEMRI) Scientific and Ethics Review Unit (SERU).

## Ethical Approvals

This study received ethical approval from the Kenya Medical Research Institute (KEMRI) Scientific and Ethics Review Unit (SERU), the Institutional Scientific and Ethics Review Committee of JOOTRH, and the Washington University in St. Louis Institutional Review Board. This study is reported in accordance with the Strengthening the Reporting of Observational Studies in Epidemiology (STROBE) Statement checklist (See Appendix 2, page 2).

## RESULTS

### Sample characteristics

Between 2014 and 2024, a total of 12,657 cancer records were identified from JOOTRH patient registries. After excluding participants aged <18, duplicates, empty patient files, incomplete medical records, non-cancer diagnoses, and unconfirmed cancer cases, 3,978 records were included in the final analytical sample (Figure 1).

[Figure 1: Study Flow Diagram (N=3978)]

The majority of cancer cases, 64% (n= 2,564), were female, and the median age of the sample was 54 years (IQR: 42.0, 65.1) (Table 1, Figure 2).

**Table 1:** Baseline demographic and clinical characteristics of cancer cases, by gender.

|  | Male (N=1414) | Female (N=2564) | Total (N=3978) | p value |
| --- | --- | --- | --- | --- |
| **Age, year** |  |  |  | <0.0001<sup>1</sup> |
| Median, IQR | 59.6 (45.5, 70.6) | 50.4 (40.0, 62.1) | 53.9 (42.0, 65.1) |  |
| **Age group, years** |  |  |  | <0.0001<sup>1</sup> |
| 18–25 | 67 (53.2%) | 59 (46.8%) | 126 (3.2%) |  |
| 26–35 | 92 (23.1%) | 307 (76.9%) | 399 (10.0%) |  |
| 36–45 | 183 (24.7%) | 559 (75.3%) | 742 (18.7%) |  |
| 46–65 | 565 (33.1%) | 1144 (66.9%) | 1709 (43.0%) |  |
| ≥66 | 507 (50.6%) | 495 (49.4%) | 1002 (25.2%) |  |
| **HIV status, n (%)** |  |  |  | <0.0001<sup>1</sup> |
| Positive | 339 (26.0%) | 966 (74.0%) | 1305 (32.8%) |  |
| Negative | 777 (41.5%) | 1094 (58.5%) | 1871 (47.0%) |  |
| Unknown | 298 (37.2%) | 504 (62.8%) | 802 (20.2%) |  |
| **Highest formal schooling, n (%)** |  |  |  | <0.0001<sup>1</sup> |
| None | 138 (40.6%) | 202 (59.4%) | 340 (8.5%) |  |
| Primary school (1st–8th class) | 105 (33.4%) | 209 (66.6%) | 314 (7.9%) |  |
| Secondary/High school | 98 (43.6%) | 127 (56.4%) | 225 (5.7%) |  |
| Post-secondary | 105 (56.5%) | 81 (43.5%) | 186 (4.7%) |  |
| Unknown | 968 (33.2%) | 1945 (66.8%) | 2913 (73.2%) |  |
| **Employment status, n (%)** |  |  |  | 0.0080<sup>2</sup> |
| Employed | 790 (37.3%) | 1327 (62.7%) | 2117 (53.2%) |  |
| Not employed | 119 (29.6%) | 283 (70.4%) | 402 (10.1%) |  |
| Unknown | 505 (34.6%) | 954 (65.4%) | 1459 (36.7%) |  |
| **Marital status, n (%)** |  |  |  | <0.0001<sup>2</sup> |
| Married | 1047 (42.6%) | 1411 (57.4%) | 2458 (61.8%) |  |
| Single | 93 (40.8%) | 135 (59.2%) | 228 (5.7%) |  |
| Divorced | 25 (32.5%) | 52 (67.5%) | 77 (1.9%) |  |
| Unknown | 174 (38.8%) | 275 (61.2%) | 449 (11.3%) |  |
| Widowed/Widower | 75 (9.8%) | 691 (90.2%) | 766 (19.3%) |  |
| **Payment method, n (%)** |  |  |  | 0.0825<sup>2</sup> |
| Cash | 369 (36.4%) | 644 (63.6%) | 1013 (25.5%) |  |
| Insurance | 829 (36.3%) | 1457 (63.7%) | 2286 (57.5%) |  |
| Unknown | 216 (31.8%) | 463 (68.2%) | 679 (17.1%) |  |
| **Smoking history, n (%)** |  |  |  | <0.0001<sup>2</sup> |
| Never | 546 (33.7%) | 1072 (66.3%) | 1618 (40.7%) |  |
| Unknown | 679 (32.4%) | 1417 (67.6%) | 2096 (52.7%) |  |
| Ever | 189 (71.6%) | 75 (28.4%) | 264 (6.6%) |  |
| **Alcohol use history, n (%)** |  |  |  | <0.0001<sup>2</sup> |
| Never | 464 (31.0%) | 1032 (69.0%) | 1496 (37.6%) |  |
| Unknown | 690 (32.7%) | 1420 (67.3%) | 2110 (53.0%) |  |
| Ever | 260 (69.9%) | 112 (30.1%) | 372 (9.4%) |  |
| **Any comorbidity, n (%)** |  |  |  | 0.0012<sup>2</sup> |
| No comorbidities | 1248 (34.8%) | 2343 (65.2%) | 3591 (90.3%) |  |
| ≥1 comorbidity | 166 (42.9%) | 221 (57.1%) | 387 (9.7%) |  |
| **Family history of cancer, n (%)** |  |  |  | 0.9192<sup>2</sup> |
| Yes | 39 (35.8%) | 70 (64.2%) | 109 (2.7%) |  |
| No | 472 (36.0%) | 840 (64.0%) | 1312 (33.0%) |  |
| Unknown | 903 (35.3%) | 1654 (64.7%) | 2557 (64.3%) |  |
| **Cancer Types, n (%)** |  |  |  | <0.0001<sup>2</sup> |
| Cervical cancer | 0 (0.0%) | 961 (100.0%) | 961 (24.2%) |  |
| Esophageal cancer | 271 (50.0%) | 271 (50.0%) | 542 (13.6%) |  |
| Breast cancer | 16 (3.3%) | 468 (96.7%) | 484 (12.2%) |  |
| Prostate cancer | 321 (100.0%) | 0 (0.0%) | 321 (8.1%) |  |
| Colorectal cancer | 82 (46.6%) | 94 (53.4%) | 176 (4.4%) |  |
| Non-Hodgkin lymphoma | 57 (53.3%) | 50 (46.7%) | 107 (2.7%) |  |
| Head and neck cancer | 70 (63.6%) | 40 (36.4%) | 110 (2.8%) |  |
| Kaposi sarcoma | 81 (69.2%) | 36 (30.8%) | 117 (2.9%) |  |
| Ovarian cancer | 0 (0.0%) | 94 (100.0%) | 94 (2.4%) |  |
| Liver cancer | 53 (58.2%) | 38 (41.8%) | 91 (2.3%) |  |
| Other* | 463 (47.5%) | 512 (52.5%) | 975 (24.5%) |  |
| **Cancer stage, n (%)** |  |  |  | <0.0001<sup>2</sup> |
| Stage 1 | 10 (7.9%) | 116 (92.1%) | 126 (3.2%) |  |
| Stage 2 | 49 (17.4%) | 232 (82.6%) | 281 (7.1%) |  |
| Stage 3 | 74 (15.8%) | 393 (84.2%) | 467 (11.7%) |  |
| Stage 4 | 228 (40.2%) | 339 (59.8%) | 567 (14.3%) |  |
| Missing/unstageable by standard criteria | 1053 (41.5%) | 1484 (58.5%) | 2537 (63.8%) |  |
| **Performance status (ECOG), n (%)** |  |  |  | 0.3574<sup>2</sup> |
| 0–1 | 383 (36.4%) | 668 (63.6%) | 1051 (26.4%) |  |
| 2–4 | 75 (31.5%) | 163 (68.5%) | 238 (6.0%) |  |
| Unknown | 956 (35.6%) | 1733 (64.4%) | 2689 (67.6%) |  |
| **Cancer-directed treatment, n (%)** |  |  |  | 0.0001<sup>2</sup> |
| Not treated | 655 (32.7%) | 1350 (67.3%) | 2005 (50.4%) |  |
| Treated | 759 (38.5%) | 1214 (61.5%) | 1973 (49.6%) |  |
| **Treatment modality, n (%)** |  |  |  | <0.0001<sup>2</sup> |
| None | 646 (32.7%) | 1328 (67.3%) | 1974 (49.6%) |  |
| Chemotherapy alone | 479 (44.3%) | 602 (55.7%) | 1081 (27.2%) |  |
| Surgery alone | 122 (39.5%) | 187 (60.5%) | 309 (7.8%) |  |
| Radiation alone | 5 (14.7%) | 29 (85.3%) | 34 (0.9%) |  |
| Combined chemotherapy + ** | 150 (27.7%) | 391 (72.3%) | 541 (13.6%) |  |
| Other | 3(35.7%) | 5(62.5%) | 8(0.2%) |  |
| Unknown | 9(29.0%) | 22(71.0%) | 31(0.8%) |  |
| **Time from diagnosis to treatment** |  |  |  | 0.0041<sup>1</sup> |
| Median in months (IQR) | 1.6 (0.5–5.5) | 1.5 (0.3–4.0) | 1.5 (0.4–4.6) |  |
| N (missing) | 626 (788) | 954 (1610) | 1580 (2398) |  |

Footnotes: ECOG, Eastern Cooperative Oncology Group; IQR, interquartile range;
<sup>1</sup> $\chi^2$ test p-value. <sup>2</sup> Mann–Whitney U test. p-value
\* Other cancers: endometrial (uterine) cancer; penile cancer; vulvar cancer; vaginal cancer; pancreatic cancer; cholangiocarcinoma; gallbladder cancer; renal (kidney) cancer; bladder cancer; urethral cancer; thyroid cancer; melanoma; skin cancers (including squamous cell carcinoma of the skin); eye cancers brain/CNS tumors; neuroblastoma; multiple myeloma/plasma cell neoplasms; leukemias (including CML, CLL, ALL, AML, and other chronic myeloproliferative neoplasms); thymoma/thymic carcinoma; small intestine cancer; appendix cancer; primary peritoneal cancer; gestational trophoblastic neoplasms (including choriocarcinoma); germ cell tumors (including testicular and extracranial germ cell tumors); Wilms tumor/nephroblastoma; sarcomas (including soft tissue and bone sarcoma subtypes); carcinoma of unknown primary; and metastatic cancer.
\*\* Includes: chemotherapy + surgery; chemotherapy + radiation; chemotherapy + surgery + radiation

[Table 1]

[Figure 2: Distribution of cancer cases by year with percent female]

Almost one third, 33% (n=1305) of patients with cancer were people living with HIV, 47% (n=1871) were HIV negative, and 20% (n=802) had unknown HIV status (Table 1, Figure 3). The proportion of cancer patients with HIV was highest among those with penile cancer (85%, 61/72), Kaposi sarcoma (82%, 96/117), and cervical cancer (53%, 509/961).

[Figure 3: Distribution of top 10 cancer diagnoses by HIV status]

Two-thirds of cancer cases had no staging data, and among those with relevant data, late-stage presentation (stage III-IV) was predominant at 72% (n=1034) (Table 1). Late-stage presentation was highest for patients with cervical cancer (35%), followed by breast cancer (15%) and prostate cancer (11%). Half of cancer cases were not on cancer-directed treatment, and functional status was assessed using the Eastern Cooperative Oncology Group (ECOG) performance scale.^19^ A substantial proportion of participants had unknown ECOG status (67.6%, n= 2689). Overall, 26.4% (n=1,051) had an ECOG performance status of 0-1, while 6.0% (n=238) had an ECOG performance of ≥2 at presentation.

### Trends in cancer cases

The volume of cancer cases at the clinic increased over time from 29 in 2014 to 708 in 2024 (Figures 2 & 4, Appendix Table A1). From 2014 to 2024, the overall average case counts increased by 38% annually (95% CI: 29–48) (Appendix Table A2). Cancer cases among females exceeded those among males throughout. The five most common cancer diagnoses were cervical cancer (24%, n=961), esophageal cancer (13.6%, n=542), breast cancer (12%, n=484), prostate cancer (8%, n=321), and colorectal cancer (4%, n=176) (Table 1, Figure 4). Cervical cancer showed the most pronounced average annual increase at 50.8% (95% CI: 28% - 78%), followed by prostate cancer at 37% (95% CI:28–47) (Figure 4, Appendix Table A2). [Figure 4: Annual trends in percentage distribution of the top 10 cancers from 2014-2024]

### Clinical outcomes: Clinical status and survival outcomes

The median follow-up time in the sample was 1.1 months (IQR: 0.1 – 7.2) (Table 2), reflecting early censoring in this cohort (Appendix Table A3). At the end of follow-up, there were 723 deaths (18%), 12 of which did not have a confirmed date of death (2%). Loss to follow-up was high, with 51% of patients having a lapse in clinic visits of ≥180 days. An additional 11% were confirmed to have transferred to other facilities.

**Table 2:** Clinical outcomes.

| **Panel A: Follow-up time and clinical status, overall and by gender** |  |  |  |  |
| --- | --- | --- | --- | --- |
|  | **Male (N=1414)** | **Female (N=2564)** | **Total (N=3978)** | **p value** |
| **Follow-up time, months** |  |  |  | 0.7870<sup>2</sup> |
| Median (IQR) | 1.1 (0.0–7.3) | 1.1 (0.1–7.1) | 1.1 (0.1–7.2) |  |
| N (missing) | 1414 (0) | 2564 (0) | 3978 (0) |  |
| **Clinical status, n (%)** |  |  |  | 0.0417<sup>1</sup> |
| Alive | 298 (36.7%) | 513 (63.3%) | 811 (20.4%) |  |
| Dead* | 274 (37.9%) | 449 (62.1%) | 723 (18.2%) |  |
| Lost to follow-up | 715 (35.4%) | 1304 (64.6%) | 2019 (50.8%) |  |
| Transfer out | 127 (29.9%) | 298 (70.1%) | 425 (10.7%) |  |
| **Panel B: Survival outcomes, overall** |  |  |  |  |
|  | **1-year survival, %**<br>**(95% CI)** | **3-year survival, %**<br>**(95% CI)** | **5-year survival, %**<br>**(95% CI)** |  |
| **Overall survival (all cases)** | 72.3% (70.2–74.3) | 58.2% (54.8–61.4) | 48.3% (43.3–53.1) |  |
| **Median survival time** | 48.36 months |  |  |  |

Footnote: IQR=interquartile range. <sup>1</sup>Chi-Square p-value; <sup>2</sup>Mann-Whitney U p-value; \*includes confirmed and unconfirmed deaths; 12 were unconfirmed.

The median survival time in the sample was 48.4 months (Table 2). Three-year overall survival was 58.2% (95% CI 54.8-61.4), decreasing to 48.3% (95% CI 43.3-53.1) at 5 years.

Comparing patients who received cancer-directed treatment versus those untreated, survival was statistically significantly different: three- and five-year survival estimates among the treated were 67% and 57%, versus 32% and 16% in the untreated, respectively (Wilcoxon p<0.001) (Figure 5: Panel A & B).

Survival also varied significantly by HIV status. Patients without HIV had significantly higher five-year survival (52.3%) than those with HIV (46.3%) and those with unknown HIV status (39.1%) (P<0.001) (Figure 5: Panel A & C).

[Figure 5: Kaplan Meier Curves of survival by treatment status and HIV status]

### Factors influencing mortality risk

After adjusting for demographic, clinical, and health-system covariates (Figure 6; Appendix Table A4), receipt of cancer-directed treatment was the strongest independent protective factor (aHR, 0.27; 95% CI, 0.22 - 0.32; *P* < 0.001). Age was not significantly associated with mortality. Poor performance status (ECOG ≥2) and missing ECOG score were the strongest independent predictors of mortality (ECOG ≥2: aHR, 3.04; 2.18 - 4.24; *P* < 0.001; ECOG unknown: 2.80; 2.23 - 3.51; *P* < 0.001).

HIV-positive (aHR, 1.43; 1.19 - 1.71; *P* < 0.001) and HIV-unknown (1.48; 1.20 - 1.81; *P* < 0.001) status were independently associated with a higher mortality risk when compared to HIV negative status.

[Figure 6: Forest plot representing multivariable Cox proportional hazards model for survival]

### Sensitivity analyses

Sensitivity analyses using alternative assumptions for missing dates of death and informative censoring confirmed the robustness of the findings (Appendix Tables A5-A6, Appendix Figures A4-A5). Although some variation in 5-year survival estimates and selected covariates was observed, overall survival patterns across HIV strata and the associations between cancer-directed treatment, poor performance status, HIV status, and mortality remained consistent.

## DISCUSSION

In this retrospective cohort of patients with cancer in Western Kenya, case counts increased rapidly between 2014 and 2024, rising by 38% annually overall and by 51% annually for cervical cancer. HIV prevalence was high: 33% of cancer patients were living with HIV, while HIV status was unknown for 20% at the time of cancer diagnosis. Only half of patients received documented cancer-directed treatment, and loss to follow-up was substantial. After adjustment, receipt of cancer-directed treatment remained the strongest modifiable predictor of survival, whereas both HIV-positive and unknown HIV status were independently associated with increased mortality. Overall, our findings indicate: the cancer burden in western Kenya is rising rapidly and is deeply intertwined with the regional HIV epidemic; cancer-directed treatment is the single largest modifiable determinant of survival yet reaches only half of patients; and routine HIV testing with HIV-informed care planning at cancer diagnosis is essential for addressing cancer outcomes in this setting.

The rapid increase in cancer diagnoses suggests that demand for oncology services in Western Kenya is expanding faster than the regional health system capacity. The observed 38% annual increase likely reflects both rising cancer incidence associated with demographic and epidemiologic transitions, as well as the continued expansion of oncology services. Kenya’s National Cancer Control Plan has prioritized prevention, screening, and treatment scale-up over this period, and programs such as HPV vaccination, introduced into the national routine immunization schedule 2019, and organized cervical cancer screening are directly relevant to two of the most rapidly growing diagnoses in our cohort.^22^ Yet, the 51% average annual growth in cervical cancer and the predominance of stage III/IV disease suggest that current prevention and early-detection efforts lag behind. Our findings are consistent with reports across Africa showing that most cancers continue to present at advanced stages, contributing substantially to poor survival.^23^ Notably, the population-based SURVCAN-3 analysis demonstrated that cancer survival across the region remains substantially lower than in high-income settings, associated with late-stage presentation and constrained treatment capacity;^24^ The survival profile and late-stage presentation observed in our cohort in Western Kenya are consistent with these findings. Together, these findings indicate that reductions in cancer mortality in Western Kenya require investment in primary prevention, early detection, and in the diagnostic pathway upstream of oncology consultation, including integration with HIV testing services.

Only half of patients received documented cancer-directed treatment, and more than half were lost to follow-up, underscoring major deficiencies in treatment access and engagement. Receipt of cancer treatment was the strongest modifiable predictor of survival, reducing adjusted mortality by approximately 73%, and this finding remained robust across all sensitivity analyses. This gap reflects widespread inequities in multimodal cancer therapy across sub-Saharan Africa.^3^ Persistent barriers to cancer care across Africa include high out-of-pocket costs, limited oncology infrastructure, workforce shortages, and long travel distances to tertiary centers.^25^ Kenya’s longstanding HIV program has successfully implemented patient navigation, community health worker support, mobile phone tracing, and decentralized models of care to improve retention.^26,27^ Adapting these strategies to oncology may be one of the most feasible ways to reduce treatment interruptions and improve survival. Taken together, these findings suggest that the largest and most immediate gains in cancer survival in the high HIV prevalence western Kenya region may come from closing the treatment-access gap and building oncology retention systems.

Nearly a third of patients were living with HIV, and one-fifth had no documented HIV status at cancer diagnosis, highlighting persistent gaps in HIV ascertainment despite Kenya’s well-established HIV program. Both groups experienced significantly elevated mortality (HIV-positive aHR 1·43, 95% CI 1·19 -1·71; HIV-unknown aHR 1·48, 1·20 - 1·81), with both signals preserved across every sensitivity specification. Consistent with prior studies in Sub-Saharan Africa, the association between HIV and increased mortality may reflect multiple factors described in the literature, including HIV-related immunosuppression, interruptions or delays in HIV care and treatment, drug-drug interactions between antiretroviral and anticancer therapies, and fragmented HIV-oncology care.^14,28^ We extend previous findings by the ABC-DO cohort, which demonstrated significantly lower 3-year survival among women with HIV than among HIV-negative women with breast cancer across five SSA countries, with the disparity most pronounced in non-metastatic disease.^29^ Our findings extend previous work by demonstrating this association across multiple cancer types while also identifying unknown HIV status as an independent marker of poor prognosis. Despite Kenya having a well-established HIV testing and treatment infrastructure and evidence supporting routine opt-out HIV testing in oncology settings,^30^ a fifth of patients in our cohort entered oncology care without a documented HIV status, suggesting persistent gaps at the point of cancer diagnosis. The unknown-HIV category likely conceals a mix of undiagnosed people with HIV who are not on antiretroviral therapy, patients too late or too sick for a complete workup, and patients whose engagement with the health system is too limited for baseline testing, each with prior reasons for worse outcomes. These findings suggest that, in high-HIV-prevalence settings, the HIV epidemic remains materially entangled with cancer outcomes, and that routine opt-out HIV testing at oncology entry is a necessary first step towards HIV-informed, integrated cancer care.

Our findings have implications for cancer policy in Kenya and comparable high-HIV-prevalence settings. Foremost, closing the treatment-access gap that leaves half of cancer patients untreated will require coordinated investment in the diagnostic pathway upstream of oncology, in the specialist workforce, and in treatment infrastructure. Kenya’s HIV testing and treatment infrastructure offers a pragmatic model for two related priorities: routine opt-out HIV testing at the first oncology visit and the adaptation of HIV-care retention strategies, such as patient navigation, community health worker linkage, and mobile phone-supported tracing, to reduce loss to follow-up in cancer care. Workflow improvements to support systematic documentation of performance status, cancer stage, and HIV status are an inexpensive but under-recognized solution. Future research should establish cohorts with HIV-specific covariates to better understand the HIV mortality signal and mechanisms underlying the observed survival disparities; implement studies evaluating patient navigation, quality improvement initiatives, decentralized treatment models, and integrated cancer screening within HIV care programs; and analyze existing hospital cohorts to further characterize cancer epidemiology across more regions in Kenya.

This study has notable strengths and important limitations. Its main strengths are its scale (nearly 4000 patients over a decade), comprehensive covariate assessment, and the robustness of the principal findings to two independent sensitivity analyses that test extremes of informative censoring and alternative event-date imputations. As a retrospective single-center cohort assembled from a referral-hospital registry, however, our estimates cannot be interpreted as population-level rates and may not generalize beyond western Kenya. The median follow-up was short, limiting inference about long-term survival despite the sensitivity analyses. Missing data were substantial, consistent with incomplete clinical workup in busy oncology settings, but constraining adjustment for confounding. Treatment was modeled as a fixed covariate from diagnosis. This may result in immortal time bias because patients had to survive long enough to initiate treatment, and many early deaths could not be ascertained, given the high loss to follow-up. As such, mortality is likely underestimated despite sensitivity analyses. Cancer-specific survival could not be analyzed because cause-of-death data were not available. HIV-specific covariates, including viral load and CD4 count, were unavailable, so the HIV mortality signal aggregates a heterogeneous group. The unknown-HIV finding likely reflects a mix of undiagnosed HIV, patients too sick for complete workup, and structurally disengaged patients, and should be interpreted as evidence of incomplete HIV ascertainment rather than a causal effect of limited testing. The ten-year study window also spans policy and service changes we could not adjust for analytically.

## CONCLUSION

In this decade-long cohort of cancer patients in western Kenya, survival was primarily determined by access to cancer-directed treatment and ascertainment of HIV status at cancer diagnosis. Despite substantial investments in HIV care, one in five patients entered oncology care without a documented HIV status, and only half received cancer treatment. These findings highlight two immediately actionable priorities for high-HIV-prevalence settings: closing the treatment-access gap through investments in oncology systems and integrating routine HIV testing and proven HIV-care retention strategies into cancer services.

## Acknowledgements

We thank the patients and staff of JOOTRH Oncology clinic.

We thank the Alvin J. Siteman Cancer Center at Washington University School of Medicine and Barnes-Jewish Hospital in St. Louis, MO., for the use of the Biostatistics and Qualitative Research Shared Resource, which provided biostatistical analysis service. The Siteman Cancer Center is supported in part by an NCI Cancer Center Support Grant #P30 CA091842.

We also acknowledge support from the Washington University Institute of Clinical and Translational Services (ICTS) through Just-in-Time (JIT) funding and the Midwest Development Center for AIDS Research (D-CFAR).

## Author Contributions

Conceptualization: TAO. Funding acquisition: TAO. Scientific oversight and supervision: TAO, HFA. Acquisition and curation of data: TAO, DM, HFA, DA, FA, CW, and AM. Clinical adjudication of cancer diagnoses: FA and TAO. Analysis plan: DM, BB, and TAO. Statistical analysis, directly accessed and verified the data: BB, DM, and HFA. Writing, original draft: DM and TAO. Writing, major revision, and finalization: HFA and TAO.

All authors had access to the underlying data, contributed to the interpretation of findings, critically reviewed the manuscript for important intellectual content, approved the final version, and agree to be accountable for all aspects of the work.

## Data Sharing Statement

De-identified individual participant data for this study will be made available upon request to the corresponding author, subject to approval by the Kenya Medical Research Institute (KEMRI) Scientific and Ethics Review Unit.

## Data Reflexivity Statement

This study was conducted through a partnership between investigators based in Kenya and the USA. The research team included clinicians and researchers affiliated with Jaramogi Oginga Odinga Teaching and Referral Hospital (JOOTRH) and the Kenya Medical Research Institute in Kenya, as well as Washington University in St. Louis in the USA. Kenyan investigators contributed to data acquisition and curation, clinical adjudication of cancer diagnoses, interpretation of the findings, and critical review of the manuscript.

The study used routinely collected clinical data from cancer care at JOOTRH, and the interpretation of the findings was informed by the clinical and health-system context in which these data were generated. All authors had access to the underlying data, contributed to the interpretation of the findings, critically reviewed the manuscript, approved the final version, and accepted accountability for the work. We recognize the importance of equitable international research partnerships and of interpreting facility-based data within the local context in which care is delivered.

## Ethics Approval and Consent

This study was approved by the Kenya Medical Research Institute Scientific and Ethics Review Unit (SERU), the JOOTRH Institutional Scientific and Ethics Review Committee, and the Washington University in St. Louis Institutional Review Board. Given the retrospective design and reliance on routinely collected clinical records spanning ten years, the requirement for individual informed consent was waived by these review boards. All patient data were deidentified prior to analysis to protect participant confidentiality, and the study was conducted in accordance with the Declaration of Helsinki.

