## Supplementary Material for "Cancer epidemiology and survival in a high HIV prevalence region of Kenya: a 10-year retrospective cohort study"

**Table A1: Percentage distribution of cases among top 10 cancer types by type and year**

| Cancer type | 2014 | 2015 | 2016 | 2017 | 2018 | 2019 | 2020 | 2021 | 2022 | 2023 | 2024 |
|---|---|---|---|---|---|---|---|---|---|---|---|
| Cervical Cancer | 8% | 11.5% | 8% | 27.8% | 46.4% | 35.4% | 34.3% | 37% | 24.6% | 31.7% | 33% |
| Esophageal Cancer | 4% | 11.5% | 18% | 18.5% | 6.8% | 14.8% | 14.1% | 13.6% | 22.4% | 22.5% | 22.4% |
| Breast Cancer | 44% | 44.3% | 22% | 22.2% | 19.8% | 13.7% | 16.5% | 12.8% | 17.2% | 13.9% | 13.6% |
| Prostate Cancer | 12% | 11.5% | 16% | 7.4% | 5% | 8.9% | 8.8% | 11.6% | 12.9% | 11.6% | 11% |
| Colorectal Cancer | 8% | 4.9% | 4% | 11.1% | 3.6% | 3.7% | 7.4% | 7.6% | 6.3% | 4.5% | 6.3% |
| Non-Hodgkin Lymphoma | 8% | 3.3% | 8% | 5.6% | 3.2% | 5.5% | 3.4% | 4.3% | 2.6% | 4% | 1.9% |
| Head and Neck Cancer | 8% | 1.6% | 8% | 0% | 2.3% | 3.3% | 5.4% | 3.5% | 4.9% | 2.9% | 3.2% |
| Kaposi Sarcoma | 0% | 8.2% | 12% | 3.7% | 7.7% | 7% | 6.4% | 3.5% | 2.2% | 2.2% | 2.1% |
| Ovarian Cancer | 8% | 1.6% | 2% | 0% | 4.1% | 3.7% | 2.4% | 1.8% | 3.5% | 3.1% | 3.9% |
| Liver Cancer | 0% | 1.6% | 2% | 3.7% | 1.4% | 4.1% | 1.3% | 4.3% | 3.4% | 3.6% | 2.6% |

**Table A2: Trends in top 10 cancer diagnoses/Average annual percent increase in cancer diagnoses by cancer type**

| CANCER TYPE | % INCREASE/YEAR | (95% CI) |
|---|---|---|
| **Overall** | **37.7** | **(28.7–47.6)** |
| Breast | 21.7 | (14.2–29.6) |
| Head and Neck | 27.1 | (15.3–40.1) |
| Cervical | 50.8 | (27.8–77.9) |
| Colorectal | 33.7 | (22.7–45.8) |
| Kaposi sarcoma | 9.1 | (−2.9–22.6) |
| Non-Hodgkin lymphoma | 22.7 | (11.4–35.2) |
| Other cancers | 37.2 | (28.6–46.4) |
| Ovarian | 28.1 | (18.4–38.5) |
| Penile | 24.4 | (7.9–43.5) |
| Prostate | 37.0 | (27.5–47.2) |

**Table A3: Clinical outcomes and follow-up time by HIV Status (N = 3978)**

| Characteristic | HIV Positive (N=1,305) | HIV Negative (N=1,871) | Unknown (N=802) | Total (N=3,978) | P-value |
|---|---|---|---|---|---|
| **Clinical Status, n (%)** | | | | | **<0.0001** |
| Alive | 237 (29.2) | 454 (56.0) | 120 (14.8) | 811 (20.4) | |
| Dead | 259 (35.8) | 312 (43.2) | 152 (21.0) | 723 (18.2) | |
| Lost to follow-up | 651 (32.2) | 922 (45.7) | 446 (22.1) | 2019 (50.8) | |
| Transfer Out | 158 (37.2) | 183 (43.1) | 84 (19.8) | 425 (10.7) | |
| **Follow-up time** | | | | | **<0.0001** |
| Median (IQR) | 1.0 (0.1–6.1) | 1.7 (0.2–9.4) | 0.3 (0.0–3.1) | 1.1 (0.1–7.2) | |
| N (Missing) | 1305 (0) | 1871 (0) | 802 (0) | 3978 (0) | |

*Values are presented as n (%) unless otherwise indicated.*
*Continuous variables are summarized as median (IQR).*
*P-values derived from Chi-square tests for categorical variables and Kruskal–Wallis tests for continuous variables*

**Table A4. Kaplan-Meier overall survival estimates and Cox proportional hazards model results for mortality**

**Panel A: Kaplan-Meier overall survival estimates**

| | **1-year survival, % (95% CI)** | **3-year survival, % (95% CI)** | **5-year survival, % (95% CI)** | **Median survival time, months** |
|---|---|---|---|---|
| **All patients** | 72·3% (70·2–74·3) | 58·2% (54·8–61·4) | 48·3% (43·3–53·1) | 48·4 (43·9–NR) |
| **By HIV status** | | | | |
| Negative | 76·2% (73·4–79·0) | 62·9% (58·5–67·6) | 52·3% (45·8–59·7) | 7·6 |
| Positive | 70·0% (66·4–73·7) | 56·0% (50·6–61·9) | 46·3% (38·3–55·8) | 4·1 |
| Unknown | 65·0% (59·7–70·8) | 47·7% (39·7–57·4) | 39·1% (29·0–52·6) | 2·6 |
| *Wilcoxon p <0·001* | | | | |
| **By treatment status** | | | | |
| Not treated | 46·6% (41·8–52·0) | 32·4% (24·7–42·4) | 15·7% (6·1–40·8) | 0·6 (0·4–1·1) |
| Treated | 82·5% (80·4–84·7) | 67·2% (63·5–71·0) | 57·0% (51·7–62·8) | 8·5 (6·3–NR) |
| *Wilcoxon p <0·001* | | | | |

**Panel B: Univariate and multivariate Cox proportional hazards model for overall survival**

| | **Unadjusted HR (95% CI)** | **p value** | **Adjusted HR (95% CI)** | **p value** |
|---|---|---|---|---|
| **Time to treatment, per unit increase** | 0·99 (0·98–1·00) | 0·1162 | | |
| **Age group, years (ref: 18–25)** | | | | |
| 26–35 | 0·94 (0·60–1·49) | 0·7980 | 0·93 (0·56–1·53) | 0·7679 |
| 36–45 | 0·72 (0·47–1·12) | 0·1431 | 0·73 (0·44–1·19) | 0·2070 |
| 46–65 | 0·91 (0·61–1·37) | 0·6582 | 0·91 (0·57–1·48) | 0·7124 |
| ≥ 66 | 0·98 (0·65–1·50) | 0·9417 | 0·94 (0·57–1·55) | 0·8207 |

| | Unadjusted HR (95% CI) | p value | Adjusted HR (95% CI) | p value |
|---|---|---|---|---|
| **Alcohol use (ref: Never)** | | | | |
| Ever | 1·10 (0·85–1·41) | 0·4749 | 0·79 (0·55–1·14) | 0·2124 |
| Unknown | 0·90 (0·77–1·05) | 0·1983 | 0·72 (0·37–1·41) | 0·3375 |
| **Any comorbidity (ref: At least 1 comorbidity)** | | | | |
| No comorbidities | 0·87 (0·70–1·09) | 0·2318 | 0·82 (0·66–1·04) | 0·0987 |
| **Cancer diagnosis (ref: Breast Cancer)** | | | | |
| Cervical Cancer | 1·62 (1·22–2·16) | 0·0010 | 1·11 (0·82–1·50) | 0·5050 |
| Colorectal Cancer | 1·66 (1·10–2·50) | 0·0151 | 1·79 (1·17–2·75) | 0·0070 |
| Esophageal Cancer | 3·28 (2·44–4·41) | <0·0001 | 1·89 (1·36–2·62) | 0·0001 |
| Kaposi Sarcoma | 0·88 (0·46–1·65) | 0·6805 | 0·79 (0·41–1·54) | 0·4906 |
| Non-Hodgkin Lymphoma | 2·14 (1·31–3·48) | 0·0023 | 1·75 (1·05–2·90) | 0·0300 |
| Other Cancer | 2·10 (1·61–2·73) | <0·0001 | 1·73 (1·30–2·30) | 0·0002 |
| Prostate Cancer | 1·10 (0·76–1·59) | 0·6113 | 0·98 (0·64–1·50) | 0·9234 |
| **Cancer stage (ref: Not stageable)** | | | | |
| Stage 1 | 0·35 (0·19–0·64) | 0·0006 | | |
| Stage 2 | 0·26 (0·17–0·41) | <0·0001 | | |
| Stage 3 | 0·61 (0·47–0·79) | 0·0002 | | |
| Stage 4 | 0·76 (0·63–0·93) | 0·0062 | | |
| **County of residence (ref: Kisumu)** | | | | |
| Homa Bay | 0·90 (0·71–1·13) | 0·3599 | 0·99 (0·78–1·26) | 0·9314 |
| Migori | 0·62 (0·39–0·99) | 0·0455 | 0·65 (0·41–1·03) | 0·0683 |
| Other | 0·57 (0·39–0·83) | 0·0034 | 0·60 (0·41–0·88) | 0·0087 |
| Siaya | 0·92 (0·77–1·10) | 0·3433 | 0·97 (0·81–1·16) | 0·7163 |
| Vihiga | 0·87 (0·60–1·24) | 0·4338 | 0·90 (0·62–1·31) | 0·5891 |
| **Performance status (ECOG) (ref: 0–1)** | | | | |
| ≥ 2 | 3·64 (2·63–5·06) | <0·0001 | 3·04 (2·18–4·24) | <0·0001 |

| | Unadjusted HR (95% CI) | p value | Adjusted HR (95% CI) | p value |
|---|---|---|---|---|
| Unknown | 3·82 (3·07–4·75) | <0·0001 | 2·80 (2·23–3·51) | <0·0001 |
| **Educational level (ref: None)** | | | | |
| Post Primary | 1·32 (0·88–1·99) | 0·1854 | 1·43 (0·93–2·19) | 0·1010 |
| Primary school (1st–8th class) | 1·47 (1·00–2·15) | 0·0468 | 1·43 (0·97–2·11) | 0·0683 |
| University | 1·00 (0·64–1·58) | 0·9955 | 1·27 (0·79–2·03) | 0·3256 |
| Unknown | 1·23 (0·91–1·66) | 0·1754 | 1·28 (0·94–1·75) | 0·1107 |
| **Employment status (ref: Not employed)** | | | | |
| Employed | 0·82 (0·63–1·05) | 0·1200 | 0·77 (0·59–1·01) | 0·0619 |
| Unknown | 1·17 (0·90–1·51) | 0·2485 | 1·01 (0·77–1·33) | 0·9236 |
| **Gender (ref: Female)** | | | | |
| Male | 1·11 (0·95–1·29) | 0·1791 | 1·06 (0·88–1·30) | 0·5281 |
| **HIV status (ref: Negative)** | | | | |
| Positive | 1·37 (1·16–1·62) | 0·0002 | 1·43 (1·19–1·71) | 0·0001 |
| Unknown | 1·77 (1·46–2·15) | <0·0001 | 1·48 (1·20–1·81) | 0·0002 |
| **Insurance category (ref: Cash)** | | | | |
| Insurance | 0·88 (0·74–1·05) | 0·1543 | 1·15 (0·95–1·38) | 0·1441 |
| Unknown | 0·93 (0·70–1·23) | 0·6140 | 0·60 (0·45–0·80) | 0·0005 |
| **Marital status (ref: Single)** | | | | |
| Divorced | 1·04 (0·56–1·96) | 0·8933 | 0·98 (0·50–1·92) | 0·9505 |
| Married | 1·11 (0·80–1·55) | 0·5210 | 1·05 (0·71–1·56) | 0·8088 |
| Unknown | 1·27 (0·87–1·87) | 0·2209 | 1·12 (0·72–1·74) | 0·6165 |
| Widowed/Widower | 1·05 (0·73–1·51) | 0·7994 | 0·83 (0·53–1·29) | 0·4000 |
| **Smoking status (ref: Never)** | | | | |
| Ever | 1·35 (1·01–1·79) | 0·0398 | 1·36 (0·91–2·03) | 0·1303 |
| Unknown | 0·93 (0·80–1·09) | 0·3624 | 1·15 (0·59–2·23) | 0·6845 |
| **Treatment status (ref: Not treated)** | | | | |

| | Unadjusted HR (95% CI) | p value | Adjusted HR (95% CI) | p value |
|---|---|---|---|---|
| Treated | 0·23 (0·19–0·27) | <0·0001 | 0·27 (0·22–0·32) | <0·0001 |

*Median survival time: 48·4 months.

HR=hazard ratio. CI=confidence interval. ECOG, Eastern Cooperative Oncology Group; HIV, human immunodeficiency virus.
The multivariable model is mutually adjusted for all listed covariates (excluding time to treatment).

**Table A5. Sensitivity analysis of Kaplan–Meier overall survival by HIV status under two alternative definitions of the event date**

| HIV status | Scenario 1: End of study date (AE) | | | Scenario 2: Last contact date (LC) | | |
|---|---|---|---|---|---|---|
| | 1-year survival, % (95% CI) | 3-year survival, % (95% CI) | 5-year survival, % (95% CI) | 1-year survival, % (95% CI) | 3-year survival, % (95% CI) | 5-year survival, % (95% CI) |
| Negative | 79·8% (77·1–82·3) | 64·6% (59·9–68·8) | 36·7% (29·4–44·1) | 76·2% (73·4–79·0) | 62·9% (58·5–67·6) | 52·3% (45·8–59·7) |
| Positive | 73·5% (69·8–76·8) | 58·0% (52·2–63·4) | 30·6% (22·1–39·5) | 70·0% (66·4–73·7) | 56·0% (50·6–61·9) | 46·3% (38·3–55·8) |
| Unknown | 68·6% (63·0–73·5) | 52·7% (44·0–60·6) | 34·2% (23·2–45·4) | 65·0% (59·7–70·8) | 47·7% (39·7–57·4) | 39·1% (29·0–52·6) |

*Abbreviations:* AE, administrative end-date imputation; CI, confidence interval; HIV, human immunodeficiency virus; LC, last clinic-visit imputation; NR, not reached.

**NOTE.** Estimates are derived from secondary sensitivity analyses of the main multivariable model in which patients with a missing date of death were assigned an imputed date. Under the LC (last clinic-visit) model, the date of death was imputed as the date of last clinic visit, or when unavailable as the midpoint between enrollment and study end. Under the AE (administrative end-date) model, the date of death was imputed as the study/follow-up end date. Survival probabilities are Kaplan-Meier estimates with 95% pointwise confidence intervals.

**Table A6: Sensitivity Analyses of Adjusted Hazard Ratios from Multivariable Cox Proportional Hazards Models for Overall Survival for Approach 1 and 2**

**Approach 1: Imputing the missing date of death**

A secondary sensitivity analysis of the main model was conducted whereby individuals missing a date of death were identified and the date of death imputed at the date of last clinic visit, or otherwise the midpoint between end date and enrollment (LC model). The model was re-run imputing the missing date of death as the study/follow-up end date (AE model).

| | **Main model** | | **LC model (last clinic visit)** | | **AE model (follow-up end)** | |
|---|---|---|---|---|---|---|
| **Variable** | **Adjusted HR (95% CI)** | **p value** | **Adjusted HR (95% CI)** | **p value** | **Adjusted HR (95% CI)** | **p value** |
| **Age group, years (ref: 18–25)** | | | | | | |
| 26–35 | 0·93 (0·56–1·53) | 0·7679 | 0·93 (0·56–1·53) | 0·7679 | 0·81 (0·49–1·33) | 0·3958 |
| 36–45 | 0·73 (0·44–1·19) | 0·2070 | 0·73 (0·44–1·19) | 0·2070 | 0·65 (0·40–1·07) | 0·0891 |
| 46–65 | 0·91 (0·57–1·48) | 0·7124 | 0·91 (0·57–1·48) | 0·7124 | 0·77 (0·47–1·24) | 0·2834 |
| ≥ 66 | 0·94 (0·57–1·55) | 0·8207 | 0·94 (0·57–1·55) | 0·8207 | 0·82 (0·50–1·36) | 0·4415 |
| **Any comorbidity (ref: At least 1 comorbidity)** | | | | | | |
| No comorbidities | 0·82 (0·66–1·04) | 0·0987 | 0·82 (0·66–1·04) | 0·0987 | 0·79 (0·63–0·99) | 0·0451 |
| **Cancer diagnosis (ref: Breast Cancer)** | | | | | | |
| Cervical Cancer | 1·11 (0·82–1·50) | 0·5050 | 1·11 (0·82–1·50) | 0·5050 | 1·13 (0·83–1·53) | 0·4390 |
| Colorectal Cancer | 1·79 (1·17–2·75) | 0·0070 | 1·79 (1·17–2·75) | 0·0070 | 1·87 (1·22–2·88) | 0·0041 |
| Esophageal Cancer | 1·89 (1·36–2·62) | 0·0001 | 1·89 (1·36–2·62) | 0·0001 | 2·04 (1·47–2·83) | <0·0001 |

| | | | | | | |
|---|---|---|---|---|---|---|
| Kaposi Sarcoma | 0·79 (0·41–1·54) | 0·4906 | 0·79 (0·41–1·54) | 0·4906 | 0·73 (0·37–1·45) | 0·3736 |
| Non-Hodgkin Lymphoma | 1·75 (1·05–2·90) | 0·0300 | 1·75 (1·05–2·90) | 0·0300 | 1·98 (1·19–3·29) | 0·0081 |
| Other Cancer | 1·73 (1·30–2·30) | 0·0002 | 1·73 (1·30–2·30) | 0·0002 | 1·72 (1·29–2·28) | 0·0002 |
| Prostate Cancer | 0·98 (0·64–1·50) | 0·9234 | 0·98 (0·64–1·50) | 0·9234 | 0·86 (0·56–1·32) | 0·4991 |
| **Performance status (ECOG) (ref: 0–1)** | | | | | | |
| ≥ 2 | 3·04 (2·18–4·24) | <0·0001 | 3·04 (2·18–4·24) | <0·0001 | 3·20 (2·29–4·46) | <0·0001 |
| Unknown | 2·80 (2·23–3·51) | <0·0001 | 2·80 (2·23–3·51) | <0·0001 | 2·83 (2·25–3·54) | <0·0001 |
| **Highest educational level (ref: None)** | | | | | | |
| Post Primary | _ | _ | 1·43 (0·93–2·19) | 0·1010 | 1·51 (0·99–2·33) | 0·0582 |
| Primary school (1st–8th class) | _ | _ | 1·43 (0·97–2·11) | 0·0683 | 1·46 (0·99–2·15) | 0·0540 |
| University | _ | _ | 1·27 (0·79–2·03) | 0·3256 | 1·11 (0·69–1·78) | 0·6632 |
| Unknown | _ | _ | 1·28 (0·94–1·75) | 0·1107 | 1·34 (0·98–1·82) | 0·0650 |
| **Employment status (ref: Not employed)** | | | | | | |
| Employed | _ | _ | 0·77 (0·59–1·01) | 0·0619 | 0·79 (0·60–1·03) | 0·0802 |
| Unknown | _ | _ | 1·01 (0·77–1·33) | 0·9236 | 1·04 (0·79–1·36) | 0·7853 |
| **Gender (ref: Female)** | | | | | | |
| Male | _ | _ | 1·06 (0·88–1·30) | 0·5281 | 1·09 (0·90–1·33) | 0·3679 |

| | | | | | | |
|---|---|---|---|---|---|---|
| **HIV status (ref: Negative)** | | | | | | |
| Positive | 1·43 (1·19–1·71) | 0·0001 | 1·43 (1·19–1·71) | 0·0001 | 1·37 (1·14–1·64) | 0·0007 |
| Unknown | 1·48 (1·20–1·81) | 0·0002 | 1·48 (1·20–1·81) | 0·0002 | 1·46 (1·19–1·80) | 0·0003 |
| **Payment mode (ref: Cash)** | | | | | | |
| Insurance | 1·15 (0·95–1·38) | 0·1441 | 1·15 (0·95–1·38) | 0·1441 | 1·18 (0·99–1·42) | 0·0704 |
| Unknown | 0·60 (0·45–0·80) | 0·0005 | 0·60 (0·45–0·80) | 0·0005 | 0·67 (0·50–0·90) | 0·0073 |
| **Marital status (ref: Single)** | | | | | | |
| Divorced | _ | _ | 0·98 (0·50–1·92) | 0·9505 | 1·19 (0·61–2·33) | 0·6126 |
| Married | _ | _ | 1·05 (0·71–1·56) | 0·8088 | 1·06 (0·71–1·57) | 0·7736 |
| Unknown | _ | _ | 1·12 (0·72–1·74) | 0·6165 | 1·10 (0·70–1·71) | 0·6846 |
| Widowed/Widower | _ | _ | 0·83 (0·53–1·29) | 0·4000 | 0·87 (0·56–1·35) | 0·5330 |
| **Cancer-directed treatment (ref: Untreated)** | | | | | | |
| Treated | 0·27 (0·22–0·32) | <0·0001 | 0·27 (0·22–0·32) | <0·0001 | 0·26 (0·21–0·30) | <0·0001 |

Breast Cancer is the reference category for cancer diagnosis in all models.

HR=hazard ratio. CI=confidence interval. ECOG=Eastern Cooperative Oncology Group. HIV=human immunodeficiency virus. All multivariable models are mutually adjusted for the listed covariates

**Approach 2: Worst and best-case scenarios**

Sensitivity analyses on the main multivariate model tested the extremes of informative censoring for individuals transferred out or lost to follow-up: outcome set as dead on the last date before transferring out or loss to follow-up (E0, worst-case scenario), and administrative censoring such that these individuals do not experience the outcome (E1, best-case scenario).

| | **Main model** | | **E0, worst-case** | | **E1, best-case** | |
|---|---|---|---|---|---|---|
| **Variable** | **Adjusted HR (95% CI)** | **p value** | **Adjusted HR (95% CI)** | **p value** | **Adjusted HR (95% CI)** | **p value** |
| Age group, years (ref: 18–25) | | | | | | |
| 26–35 | 0·93 (0·56–1·53) | 0·7679 | 0·87 (0·53–1·42) | 0·5729 | _ | _ |
| 36–45 | 0·73 (0·44–1·19) | 0·2070 | 0·71 (0·44–1·16) | 0·1749 | _ | _ |
| 46–65 | 0·91 (0·57–1·48) | 0·7124 | 0·86 (0·54–1·37) | 0·5246 | _ | _ |
| ≥ 66 | 0·94 (0·57–1·55) | 0·8207 | 0·84 (0·52–1·37) | 0·4935 | _ | _ |
| **Any comorbidity (ref: At least 1 comorbidity)** | | | | | | |
| No comorbidities | 0·82 (0·66–1·04) | 0·0987 | 0·72 (0·58–0·91) | 0·0054 | 0·92 (0·76–1·12) | 0·4181 |
| **Cancer diagnosis (ref: Breast Cancer)** | | | | | | |
| Cervical Cancer | 1·11 (0·82–1·50) | 0·5050 | 0·83 (0·61–1·12) | 0·2204 | 1·81 (1·41–2·33) | <0·0001 |
| Colorectal Cancer | 1·79 (1·17–2·75) | 0·0070 | 1·52 (0·99–2·33) | 0·0525 | 1·81 (1·26–2·61) | 0·0014 |
| Esophageal Cancer | 1·89 (1·36–2·62) | 0·0001 | 1·36 (0·98–1·89) | 0·0626 | 2·31 (1·75–3·05) | <0·0001 |

| | | | | | | |
|---|---|---|---|---|---|---|
| Kaposi Sarcoma | 0·79 (0·41–1·54) | 0·4906 | 0·72 (0·37–1·39) | 0·3257 | 0·99 (0·57–1·70) | 0·9659 |
| Non-Hodgkin Lymphoma | 1·75 (1·05–2·90) | 0·0300 | 1·37 (0·83–2·28) | 0·2191 | 1·82 (1·19–2·79) | 0·0061 |
| Other Cancer | 1·73 (1·30–2·30) | 0·0002 | 1·43 (1·07–1·90) | 0·0145 | 1·86 (1·45–2·39) | <0·0001 |
| Prostate Cancer | 0·98 (0·64–1·50) | 0·9234 | 0·98 (0·64–1·49) | 0·9256 | 1·13 (0·79–1·61) | 0·5068 |
| **Performance status (ECOG) (ref: 0–1)** | | | | | | |
| ≥ 2 | 3·04 (2·18–4·24) | <0·0001 | 3·00 (2·16–4·19) | <0·0001 | 1·74 (1·33–2·27) | <0·0001 |
| Unknown | 2·80 (2·23–3·51) | <0·0001 | 2·73 (2·18–3·42) | <0·0001 | 1·67 (1·43–1·96) | <0·0001 |
| **Highest educational level (ref: None)** | | | | | | |
| Post Primary | 1·43 (0·93–2·19) | 0·1010 | 1·90 (1·24–2·91) | 0·0033 | 0·88 (0·64–1·22) | 0·4374 |
| Primary school (1st–8th class) | 1·43 (0·97–2·11) | 0·0683 | 1·96 (1·33–2·89) | 0·0007 | 0·88 (0·66–1·17) | 0·3682 |
| University | 1·27 (0·79–2·03) | 0·3256 | 1·45 (0·91–2·32) | 0·1194 | 0·97 (0·69–1·38) | 0·8736 |
| Unknown | 1·28 (0·94–1·75) | 0·1107 | 1·60 (1·18–2·18) | 0·0029 | 0·85 (0·69–1·05) | 0·1394 |
| **Employment status (ref: Not employed)** | | | | | | |
| Employed | 0·77 (0·59–1·01) | 0·0619 | 0·76 (0·58–0·99) | 0·0429 | 0·88 (0·71–1·08) | 0·2162 |
| Unknown | 1·01 (0·77–1·33) | 0·9236 | 1·10 (0·84–1·44) | 0·4786 | 0·95 (0·77–1·17) | 0·6241 |
| **Gender (ref: Female)** | | | | | | |
| Male | 1·06 (0·88–1·30) | 0·5281 | 1·02 (0·84–1·24) | 0·8391 | 1·07 (0·91–1·26) | 0·4088 |

| | | | | | | |
|---|---|---|---|---|---|---|
| **HIV status (ref: Negative)** | | | | | | |
| Positive | 1·43 (1·19–1·71) | 0·0001 | 1·34 (1·11–1·60) | 0·0017 | 1·27 (1·10–1·47) | 0·0010 |
| Unknown | 1·48 (1·20–1·81) | 0·0002 | 1·36 (1·11–1·67) | 0·0034 | 1·42 (1·21–1·68) | <0·0001 |
| **Insurance category (ref: Cash)** | | | | | | |
| Insurance | 1·15 (0·95–1·38) | 0·1441 | 1·29 (1·08–1·56) | 0·0057 | 1·03 (0·89–1·19) | 0·6929 |
| Unknown | 0·60 (0·45–0·80) | 0·0005 | 0·55 (0·41–0·73) | <0·0001 | 0·81 (0·66–0·99) | 0·0365 |
| **Marital status (ref: Single)** | | | | | | |
| Divorced | 0·98 (0·50–1·92) | 0·9505 | 1·00 (0·51–1·95) | 0·9975 | 0·93 (0·55–1·59) | 0·7986 |
| Married | 1·05 (0·71–1·56) | 0·8088 | 1·03 (0·70–1·52) | 0·8620 | 1·04 (0·77–1·40) | 0·8056 |
| Unknown | 1·12 (0·72–1·74) | 0·6165 | 1·02 (0·66–1·58) | 0·9135 | 1·21 (0·86–1·70) | 0·2739 |
| Widowed/Widower | 0·83 (0·53–1·29) | 0·4000 | 0·86 (0·56–1·33) | 0·5106 | 0·82 (0·58–1·15) | 0·2457 |
| **Cancer-directed treatment (ref: Untreated)** | | | | | | |
| Treated | 0·27 (0·22–0·32) | <0·0001 | 0·44 (0·37–0·52) | <0·0001 | 0·23 (0·20–0·27) | <0·0001 |

Breast Cancer is the reference category for cancer diagnosis in all models.

HR=hazard ratio. CI=confidence interval. ECOG=Eastern Cooperative Oncology Group. HIV=human immunodeficiency virus. All multivariable models are mutually adjusted for the listed covariates.

Cancer epidemiology and survival in a high HIV prevalence region of Kenya: a 10-year retrospective cohort study
SUPPLEMENTARY MATERIAL

### Figure A4: Forest plots displaying adjusted hazard ratios for sensitivity analyses for cox model with imputation for missing date of death

**Sensitivity analysis: imputation of missing date of death (Approach 1)**

Adjusted hazard ratios from the main multivariable Cox model compared with two imputation approaches for missing date of death. Dashed vertical line indicates HR=1 (no effect).

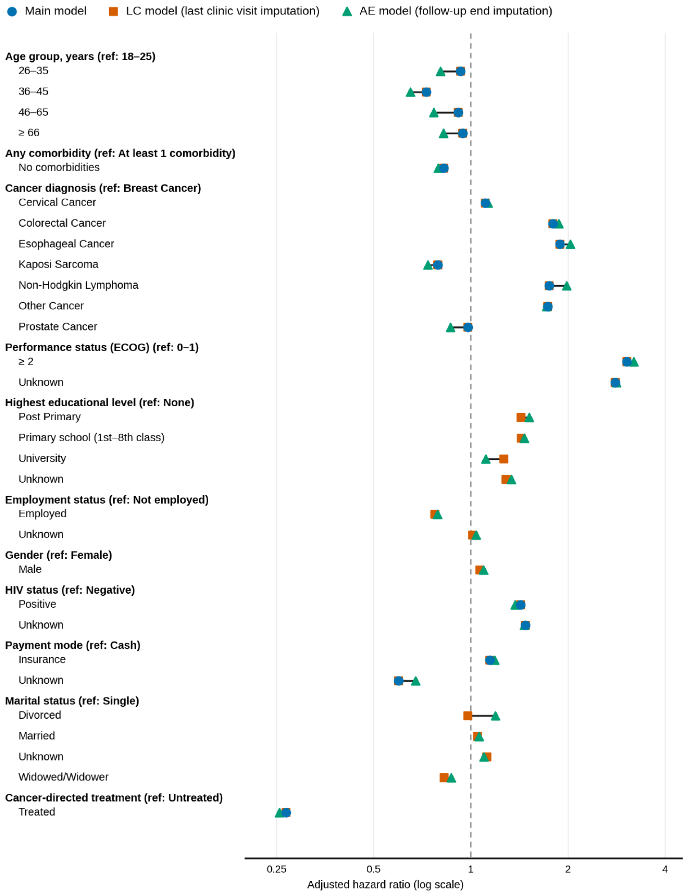


HR=hazard ratio. ECOG=Eastern Cooperative Oncology Group. HIV=human immunodeficiency virus. Breast Cancer is the reference category for cancer diagnosis in all models.

### Figure A5: Forest plots displaying adjusted hazard ratios for sensitivity analyses for the Cox model with imputation for informative censoring to mimic best and worst-case scenarios

**Sensitivity analysis: worst and best case censoring scenarios (Approach 2)**

Adjusted hazard ratios from the main multivariable Cox model compared with worst-case (E0) and best-case (E1) assumptions for informative censoring among individuals transferred out or lost to follow-up. Dashed vertical line indicates HR=1 (no effect).

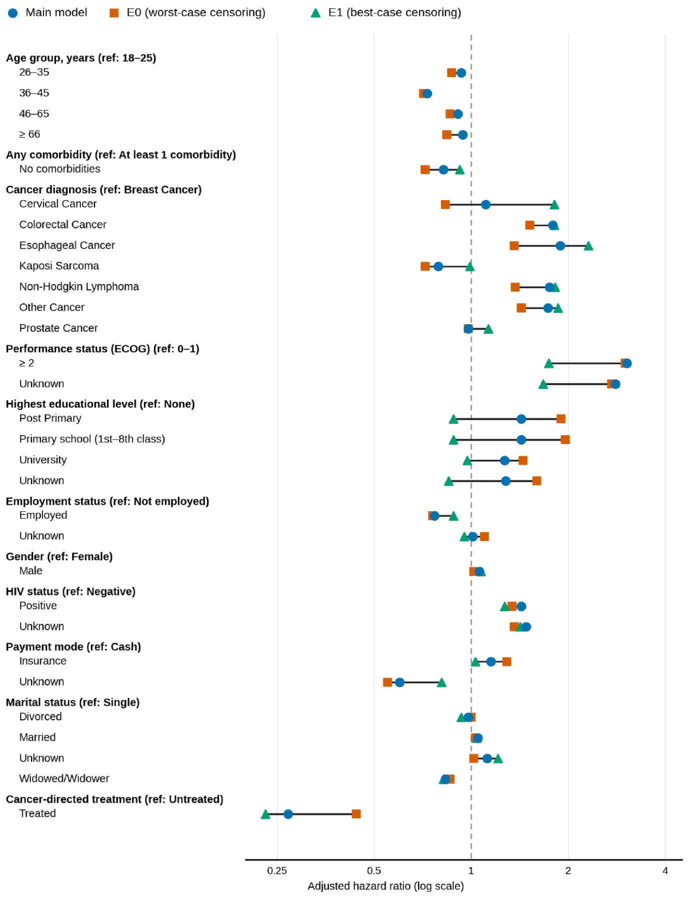


HR=hazard ratio. ECOG=Eastern Cooperative Oncology Group. HIV=human immunodeficiency virus. Breast Cancer is the reference category for cancer diagnosis in all models.
